# Polymorphism of the glucocorticoid receptor gene (NR3C1) in chronic illnesses: a protocol of a scoping review

**DOI:** 10.1101/2020.07.31.20166108

**Authors:** Y Sguassero, G Gallucci, O Bottasso

## Abstract

During process like local or systemic homeostatic disturbances due to infection, tissue injury, trauma or surgery, neoplastic growth or immunological disorders, the host responds with a defensive reaction, the acute phase response, which encompasses alterations in immune, metabolic, neuroendocrine functions. Beyond the triggering stimulus, innate immune cells, quickly manage to deal with such a threat and secrete proinflammatory cytokines like Tumour Necrosis Factor alpha (TNF-α), Interleukin 1 beta (IL-1β), and Interleukin 6 (IL-6). Beyond their immunological effects, these cytokines also activate the stress system within the central nervous system leading to the activation of the hypothalamic-pituitary-adrenal axis as an effectors arm. As such, the immune response mounted against pathogens is paralleled by a significantly altered hormonal response. In relation to the immunomodulatory influences of adrenal steroids, glucocorticoids (GCs) suppress the immune system at several levels, avoiding the possible adverse effects of an excessive immune response and helping to terminate it once the noxious stimulus was eliminated. Under some circumstances, particularly at the beginning of the immune response, GCs can also exert pro-inflammatory effects. But at high concentrations, GCs generally suppress immune and inflammatory responses arising during activation of innate and adaptive immune responses. As in many physiological processes, individual sensitivity to GCs is variable being determined by genetic and acquired factors. This kind of versatility of GCs effects, that also extends to immune cells, may be due to a series of mechanisms, including a familiar resistance linked to inactivating mutations of GR or SNP -Single Nucleotide Polymorphism-functions affecting transcription. In this context, we will conduct a Scoping Review (ScR) of the literature about SNPs in the glucocorticoid receptor gene (NR3C1).

The general aim of this ScR is to examine the extent and nature of available evidence on chronic illnesses associated with SNPs in the glucocorticoid receptor gene. The scoping review will assess evidence from all observational study designs. We will conduct a search in MEDLINE and LILACS. No language restriction will be applied. We will also search The Cochrane Library, and Epistemonikos to identify systematic reviews as an additional mechanism to identify primary studies. These searches will aim to identify all articles related to SNPs in the GR gene and will not be restricted to those evaluating specific conditions. These searches will be supplemented by scanning references of relevant retrieved articles to identify any additional relevant studies. References will be screen by titles and abstracts, remaining potentially relevant articles will be screen as full texts. Two reviewers will independently screen all identified records for relevance. Conflicts between reviewers will be solved by consensus or by the lead systematic reviewer in consultation with the content experts, if required. We will create a diagram to show the number of studies in each population condition. One reviewer will chart the data using pre-tested data-charting forms. This data charting will be double-checked for accuracy by a second reviewer. We will resolve any disagreements about data extraction by referring to the study report and through discussion. Once data is charted for all included studies, we will tabulate the data for each new condition identified. Findings from the scoping reviews will be reported according to the newly developed, PRISMA for Scoping Reviews (PRISMA-ScR). When interpreting the review findings, we will make sure that the limitations of different study designs are carefully considered, though no quality assessment will be performed.

## Background and rationale

During process like local or systemic homeostatic disturbances due to infection, tissue injury, trauma or surgery, neoplastic growth or immunological disorders, the host responds with a defensive reaction, the acute phase response, which encompasses alterations in immune, metabolic, neuroendocrine functions (1,2). While these alterations are adaptive in nature and tend to be beneficial for the host at least during the early phase of the process, an excessive or prolonged response turns out to be detrimental favouring the development of pathology and the ensuing clinical manifestations.

A great variety of organisms including bacteria, viruses, parasites, and fungi can cause infection in humans. In cases wherein, defensive mechanisms are not able to cope with the infectious insult, partly because pathogens escape from immune clearance, a state of chronic infection in the form of persistent or latent infections ensues. The same happens with disease exhibiting a degenerative or autoimmune basis in which the triggering insult cannot be removed.

Inflammation can be triggered by infectious microbes endowed with pathogen-associated molecular patterns (PAMPs), recognized by pathogen recognition receptors (PRR) present in many immune cells, i.e., macrophages, monocytes, and dendritic cells, among others (3). Undoubtedly, defence against pathogenic microbes is vital; however, an inflammatory response after sterile damage and subsequent tissue repair may be just as important from an evolutionary standpoint. Inflammation in sterile injury is initiated by the same innate pattern recognition systems used to detect microbes. However, the immunostimulatory molecular patterns in sterile inflammation differ from microbial patterns and are canonically associated with damage; for which they are called damage-associated molecular patterns (DAMPs). DAMPs are released during tissue damage (3) to initiate the inflammatory response.

Beyond the triggering stimulus, innate immune cells, quickly manage to deal with such a threat and secrete proinflammatory cytokines like Tumour Necrosis Factor alpha (TNF-α), Interleukin 1 beta (IL-1β), and Interleukin 6 (IL-6) (1–5). Beyond their immunological effects, these cytokines produced at the site of inflammation also activate the stress system within the central nervous system (CNS) leading to the activation of the hypothalamic-pituitary-adrenal (HPA) axis as an effectors arm (6). As such, the immune response mounted against pathogens is paralleled by a significantly altered hormonal response, as documented in a large series of experimental and clinical studies (7,8).

Many studies point out to an influential role of the interaction between immune and endocrine systems in orchestrating an effective defence strategy against the infective insult (8,9). Endocrine and immune systems are strictly connected by multiple mutual regulatory pathways. As above stated, proinflammatory cytokines IL-1β, IL-6, TNF-α produced in response to an infectious challenge activate the HPA axis leading to the production of adrenal steroids (6). In relation to the immunomodulatory influences of adrenal steroids, glucocorticoids (GCs) suppress the immune system at several levels, avoiding the possible adverse effects of an excessive immune response and helping to terminate it once the noxious stimulus was eliminated (10,11). Under some circumstances, particularly at the beginning of the immune response, GCs can also exert pro-inflammatory effects (12). But at high concentrations, GCs generally suppress immune and inflammatory responses arising during activation of innate and adaptive immune responses. These are critical for coping with intracellular pathogens like bacteria, fungi, and parasites (cellular immunity) as well as extracellular bacteria, soluble toxins, some viruses, and multicellular parasites (humoral mechanisms). While GCs are known to inhibit both Th1 and Th2 cytokine production by activated human T cells, they predominantly affect Th1 cytokine production, shifting T cell response towards a Th2 profile (13).

As an integrated physiological circuit, the HPA axis represents a well-conserved mechanism to control/support an intense immune-inflammatory reaction as well as for the early mobilization of immune cells and their redistribution to mount an adequate defensive response. Nevertheless, when dealing with chronic infections, the immune response needs to be sustained in time, leading to a chronic inflammatory state resulting in substantial changes in immune and endocrine responses (13–15).

Under certain circumstances, the hyperactivity of the HPA axis because of a persistent inflammatory process can also lead to desensitization of the GCs in target tissues by downregulating the GC receptor -GR-(16).

The biological function of cortisol depends on its interaction with the GR formed by the GRα dimer, which is the biologically active form (10,11,17) while the GRα-GRβ heterodimer acts as a negative dominant.

The GR is critical for mediating the action of glucocorticoids and hence influencing physiologic functions essential for life. The variable effect that GR gene mutations/polymorphisms may have on glucocorticoid signal transduction indicates that alterations in GR action may have important implications for many critical biological processes, like the physiologic responses to the immune and inflammatory reaction, in addition to other basic activities, such as growth and reproduction.

As in many physiological processes, individual sensitivity to GCs is variable being determined by genetic and acquired factors (18,19). This kind of versatility of GCs effects, that also extends to immune cells, may be due to a series of mechanisms, including a familiar resistance linked to inactivating mutations of GR or SNP -Single Nucleotide Polymorphism-functions affecting transcription. Panek et al. demonstrated that a *Tth111*I polymorphism of the *NR3C1* gene is associated with the pathogenesis of chronic bronchitis leading to the development of asthma (20). On the other hand, 4 well-characterized functional polymorphisms of the GR gene (9β, ER22 / 23EK, BclI and N363S) modulating GC sensitivity may also have to do with variation in the clinical response to GCs in patients with rheumatoid arthritis or other inflammatory diseases (21–24).

In this context, a map of the evidence is needed. A scoping review (ScR) is a form of knowledge synthesis that aims to map the spectrum of evidence and gaps in a defined area or field by systematically searching, selecting, and synthesizing available evidence (25). This ScR aims to describe the alterations at the level of SNPs in the GR receptor gene in a series of chronic inflammatory diseases, including the infectious ones, of great impact in human health as well as their repercussion in disease pathology and clinical manifestations.

### Review question

Which chronic illnesses have been studied to search for an association with single nucleotide polymorphisms (SNPs) in the glucocorticoid receptor gene?

### Objectives

We will conduct a ScR of the literature about SNPs in the glucocorticoid receptor gene (NR3C1). The general aim of this ScR is to examine the extent and nature of available evidence on chronic illnesses associated with SNPs in the glucocorticoid receptor gene. This will serve as a preliminary exercise to ascertain the value of undertaking subsequent research on this topic.

The objectives of this research are:

i. To map the published literature reporting on the SNPs in the GR gene
ii. To describe the spectrum of conditions associated with any SNPs in the GR gene
iii. To summarize the available evidence about SNPs in the GR gene
iv. To highlight existing gaps in the literature

## Methods

Our approach will be guided by established scoping review methodology (25,26). See also: https://wiki.joannabriggs.org/display/MANUAL/Chapter+11%3A+Scoping+reviews

The results of this review should be reported in keeping with PRISMA guidance for scoping reviews http://www.prisma-statement.org/Extensions/ScopingReviews

### 1.1 Identifying the research question

A well accepted methodology associated with framing of questions in systematic reviews mandates carefully specifying the patient population (P), the intervention of interest (I), the comparator (C), and the outcomes of interest (O). This is known as the “PICO” approach to formulating a research question.

#### Population

Adults with a chronic illness that could be associated with SNPs in the GR gene.

A chronic illness will be defined as a disease which have one or more of the following characteristics: they are permanent, leave residual disability, are caused by no reversible (or quite difficult to handle) pathological alteration, require special training of the patient for rehabilitation, or may be expected to require a long period of supervision, observation, or care. (Dictionary of Health Services Management, 2d ed)

We will focus on:

- Infectious diseases such as tuberculosis, Chagas disease, lepra, malaria
- Autoimmune disorders such as polyarteritis nodosa, systemic lupus erythematosus, rheumatoid arthritis
- Obesity, metabolic syndrome, diabetes type II
- Allergic conditions such as asthma
- Other chronic conditions

**We will exclude patients with neoplasms or any other chronic conditions not encompassing an inflammatory or immunological component**.

#### Intervention

We will include studies assessing the following main SNPs in the GR gene (e.g. “9beta”, “exon 9beta”, “rs6198”, “ER22/23EK”, “rs6195”, “rs56149945”, “N363S”, “rs41423247”, “Bcll”, “rs2307674”, “rs6190”, “Tth111I”, “A3669G”). This list is not a comprehensive list, and any identified SNPs in the glucocorticoid receptor gene reported in studies meeting our inclusion criteria will be considered.

#### Outcomes

We will estimate the global frequency of the SNPs in the GR receptor gene, and its potential relationship with a certain chronic condition.

#### Types of studies

The scoping review will assess evidence from all observational study designs. We will also include systematic reviews and meta-analysis of observational studies.

**We will exclude commentaries, letters, and narrative reviews**.

### 1.2 Search strategy

We will conduct a search in MEDLINE and LILACS. The search will be limited to studies conducted in humans. Ongoing trials will also be searched, but not searches will be run for grey literature. No language restriction will be applied.

We will also search The Cochrane Library, and Epistemonikos to identify systematic reviews as an additional mechanism to identify primary studies.

These searches will aim to identify all articles related to SNPs in the GR gene and will not be restricted to those evaluating specific conditions.

The following search strategy will be executed:

**(Receptors, Glucocorticoid OR NR3C1) AND (Polymorphism, Genetic OR Polymorphism, Single Nucleotide)**

These searches will be supplemented by scanning references of relevant retrieved articles to identify any additional relevant studies.

### 1.3 Study selection

Duplicates will be identified and removed. The remaining references will be screen by titles and abstracts (Level 1), remaining potentially relevant articles will be screen as full texts (Level 2).

Level 1 will consist of two reviewers (GG, OB) independently screening for relevance based on title and abstract. At Level 2, full texts will be retrieved and two reviewers (GC, OB) will independently assess the article for relevancy. Conflicts between reviewers will be solved by consensus or by the lead systematic reviewer in consultation with the content experts, if required.

We will create a diagram to show the number of studies in each population condition (27).

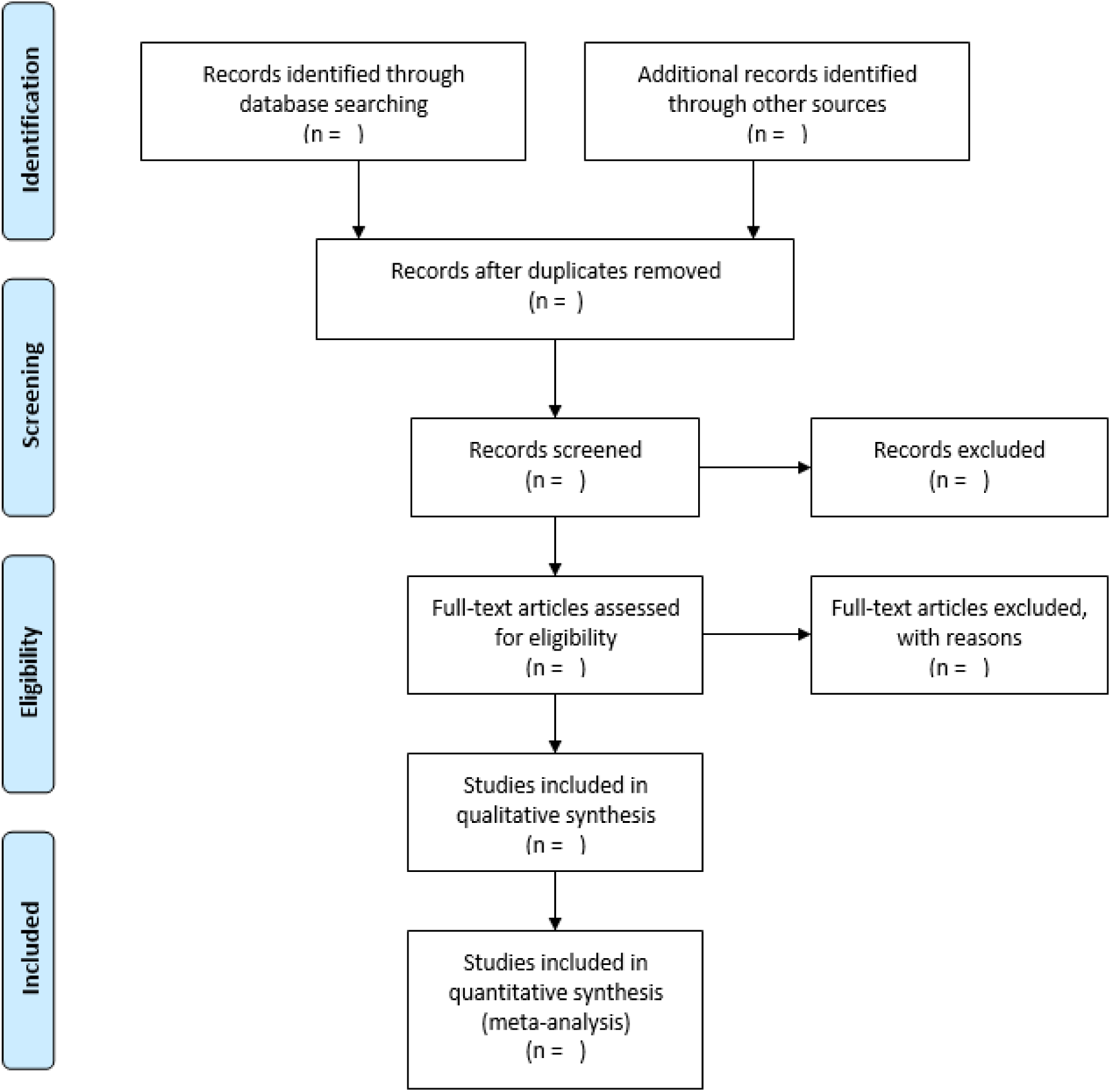

A list of sources excluded following full-text review with primary reasons for exclusion.

### 1.4 Data collection and charting

One reviewer (GC) will chart the data using pre-tested data-charting forms. This data charting will be double-checked for accuracy by a second reviewer. We will resolve any disagreements about data extraction by referring to the study report and through discussion.

Once data is charted for all included studies, we will tabulate the data for each new condition identified.

For each included study, we will chart the following information:

- Author (s)
- Year of publication
- Country
- Study design
- Number of participants
- Description of condition, i.e., type of chronic illness (as defined by authors)
- Description of population: age, gender, ethnicity.
- Description of genetic polymorphism: allele and genotype frequencies extracted or calculated from published data if possible, genotyping method, sample type,
- Glucocorticoid resistance (as measured by author, e.g. *in vivo* or *in vitro*)
- Key conclusions from the study.

### 1.5 Collating, summarizing, and reporting the results

We will include all observational study designs reporting on the polymorphism of the receptor of GC gene. Therefore, studies meeting the inclusion criteria in the scoping review may be:

- Case-control studies
- Cohort studies (prospective and retrospective)
- Cross-sectional studies
- Case series
- Case reports

Study characteristics (e.g., author, year, country, number of participants, outcomes) of the included studies will be presented in tabular form or by using visual representations (e.g., bar charts). Results may also be summarized by study design for each population/condition and presented in tables or charts as appropriate.

Findings from the scoping reviews will be reported according to the newly developed, PRISMA for Scoping Reviews (PRISMA-ScR) (28).

When interpreting the review findings, we will make sure that the limitations of different study designs are carefully considered, though no quality assessment will be performed. We anticipate that there will be differences in findings for different conditions and we will report data without aggregation to make this clear.

### Timeline

Phases for the scoping review include:

Reading about scoping reviews methodology

Protocol development and registering

Search strategy and electronic searches

Search, title & abstract screening

Full text screening

Extracting the data

Presentation of the results and data charting

PRISMA flow-chart

Report writing

## Data Availability

This is a retrospective study and aggregate data will be extracted from published papers.

## Contributions of authors

YS, contributed to methodological and technical expertise, developed the search strategy, and conducted the electronic searches. She also drafted the protocol, and coordinated contributions from all co-authors. GC contributed to develop the search strategy in MEDLINE and helped to develop the protocol. OB contributed with content expertise and helped to develop the protocol.

## Declaration of interest

Yanina Sguassero has no conflict of interest to declare.

Georgina Gallucci has received financial support for performing studies, including the conduct of this scoping review. In no situation, the funder agency had any influence on the results of the work.

Oscar Bottasso is a senior researcher from The National Council of Scientific and Technological Research and the National University of Rosario, Argentina.

